# Retrospective Observational Study on an AI-Powered Symptom Checker for Pre-Diagnosis of Anxiety Disorders

**DOI:** 10.1101/2023.05.24.23290449

**Authors:** Abel Andrés Orelogio, Oscar Garcia-Esquirol, María Eugenia Rodriguez, Andrea Malet Gausa, Ernest Sarrias Ramis

## Abstract

**Objective:** To describe the data obtained about diagnoses of anxiety disorders from the Mediktor database to reveal the relationship with the current scientific evidence.

**Methods:** This retrospective observational study uses secondary data obtained from the Mediktor database and evaluated age, sex, and the most frequent reason for consultation. Descriptive statistics were used to analyze the variables of the sample and their association with pre-diagnosis anxiety disorders. A significance level of p < 0.05 was set for all statistical tests, and all analyses were conducted using Power BI and Google Sheets.

**Results:** Young adults (18-35 years old) were more likely to be pre-diagnosed with anxiety disorders compared to individuals in other life stages and the proportion of anxiety disorders was found to be higher in females compared to males, both with a statistically significant p-value of less than 0.01.

**Conclusions:** The data observed in Mediktor relates to current scientific evidence and it has the potential as a technological tool for early detection of anxiety disorders.

## Introduction

There are numerous mental health conditions that affect the general population, but anxiety disorders (AD) are the most common ^1,2^. According to the WHO, AD are the tenth most frequent health-related impairment worldwide. There are around 264 million people around the world who suffer from these illnesses which affect people of all ages, including children, adolescents, and adults ^3,6^.

Depending on the individual symptoms, these diseases can lead to a variety of psychological problems. Excessive fear, avoidance of perceived threats, discomfort, and panic attacks are some examples of symptoms of AD^4,5^

Anxiety disorders can disrupt people’s social, occupational, and personal lives^7^. They can also result in significant healthcare expenses due to medical treatment, loss of productivity, and disability^8^. These conditions’ symptoms often co-occur with other mental health conditions, such as depression, substance abuse, and post-traumatic stress disorder^9^.

Research indicates a significant correlation between the utilization of psychoactive drugs and anxiety-related issues, with those who engage in drug use exhibiting poorer clinical outcomes^10^. While previous studies have solely identified a relationship between smoking and Panic Disorder^11^, more recent inquiries have expanded to encompass all anxiety disorders and their associations with substance use, garnering increased attention^12^.

It is critical to discover and diagnose anxiety disorders early in order for patients to receive appropriate care and therapy. However, identifying anxiety disorders can be difficult because symptoms are not always obvious or might be confused with other conditions. The use of technological tools such as Mediktor provide an objective analysis of the symptoms experienced by the patient, which facilitates rapid detection of the condition and referral to an appropriate care center.

Mediktor is a cutting-edge symptom checker that employs the most recent advances in artificial intelligence and natural language processing to assess a patient’s symptoms accurately. Unlike other symptom checkers, Mediktor is presented as an easily accessible tool that can be used by anyone. Users can receive accurate and reliable information regarding possible diagnoses by simply stating their symptoms in their own words.

The objective of the present study was to describe the data obtained about diagnoses of anxiety disorders from the Mediktor database to reveal the relationship between the data collected by Mediktor and current scientific evidence.

## Methods

This retrospective observational study uses secondary data obtained from the Mediktor database. We employed Power BI and Google Sheets to extract data. Mediktor is a symptom checker that utilizes artificial intelligence technology and contains a unique database with over 3,172,141 evaluations.

Mediktor provides a main pre-diagnosis and up to nine alternative pre-diagnoses. The main pre-diagnosis is based on the number of symptoms the user responds to and whether the combination of those symptoms is accurate for a given disease.

Evaluations were obtained from January 1, 2018 to January 1, 2023. A sample of 2,685,495 evaluations with a primary diagnosis was selected, of which 92,964 had “Anxiety Disorder” as the main pre-diagnosis and met the study’s inclusion criteria. The inclusion criteria consisted of being over six years old and having “Anxiety Disorder” as the main diagnosis.

This study evaluated age, sex, and the most frequent reason for consultation. Age was divided into the following life stages: childhood (6 to 11 years old), adolescence (12 to 17 years old), young adults (18-35 years old), middle-aged adults (36-65 years old), and older adults (> 65 years old). Sex was classified as male or female, and the top five reasons for consultation were selected and cross-referenced across different age groups.

The anonymity of the users and the confidentiality of the data provided by the evaluations were assured. All information collected from the Mediktor database was considered confidential according to established data protection and privacy regulations.

Descriptive statistics were used to analyze the variables of the sample and their association with pre-diagnosis anxiety disorders. A significance level of p < 0.05 was set for all statistical tests, and all analyses were conducted using Power BI and Google Sheets.

## Results

During the studied period, 2,892,638 evaluations were observed in Mediktor. Of these evaluations, 2,649,986 presented main pre-diagnoses, and 92,964 had an anxiety disorder as the main pre-diagnosis.

Table 1 shows the age distribution, number of evaluations with main pre-diagnoses, and their distribution by sex. Table 2 presents the same parameters but refers specifically to the main pre-diagnosis of “Anxiety Disorder.”

**Table 1:** Distribution by life stage, main prediagnoses, and sex distribution

| Life stages | Main pre-diagnosis<br>n (%) | Sex |  |  |  |
| --- | --- | --- | --- | --- | --- |
|  |  | Male |  | Female |  |
|  |  | n | % | n | % |
| Childhood | 32,003 (1.2) | 14,852 | 46.41 | 17,151 | 53.59 |
| Adolescence | 325,673 (12.29) | 89,715 | 27.55 | 235,958 | 72.45 |
| Young adults | 1,560,625 (58.89) | 435,597 | 27.91 | 1,125,028 | 72.09 |
| Middle-aged adults | 687,617 (25.95) | 244,864 | 35.61 | 442,753 | 64.39 |
| Older adults | 44,068 (1.66) | 19,732 | 44.78 | 24,336 | 55.22 |
| Total | 2,649,986 | 804,760 |  | 1,845,226 |  |

**Table 2:** Distribution by life stage, number of times prediagnosed “Anxiety Disorders” and gender distribution

| Life stages | Main pre-diagnosis | Sex |  |  |  |
| --- | --- | --- | --- | --- | --- |
|  | n (%) | Male |  | Female |  |
|  |  | n | % | n | % |
| Childhood | 419 (0.45) | 128 | 30.55 | 291 | 69.45 |
| Adolescence | 17,212 (18.51) | 3,564 | 20.71 | 13,648 | 79.29 |
| Young adults | 57,260 (61.59) | 14,451 | 25.24 | 42,809 | 74.76 |
| Middle-aged adults | 17,454 (18.78) | 5,206 | 29.83 | 12,248 | 70.17 |
| Older adults | 619 (0.67) | 185 | 29.89 | 434 | 70.11 |
| Total | 92,964 | 23,534 |  | 69,430 |  |

In this last table, we can observe that young adults were more likely to be pre-diagnosed with anxiety disorders compared to individuals in other life stages, with a statistically significant p-value of less than 0.01. Additionally, the proportion of anxiety disorders was higher in females than males, with a statistically significant p-value of less than 0.01.

The five most common symptoms leading to consultation, in descending order, were “Feeling nervous and anxious,” “Dizziness,” “Fear of having a panic attack and losing control,” “Chest pain,” and “Headache.” Figure 1 presents the different reasons for consultation and their distribution according to life stage.

**Figure 1:**
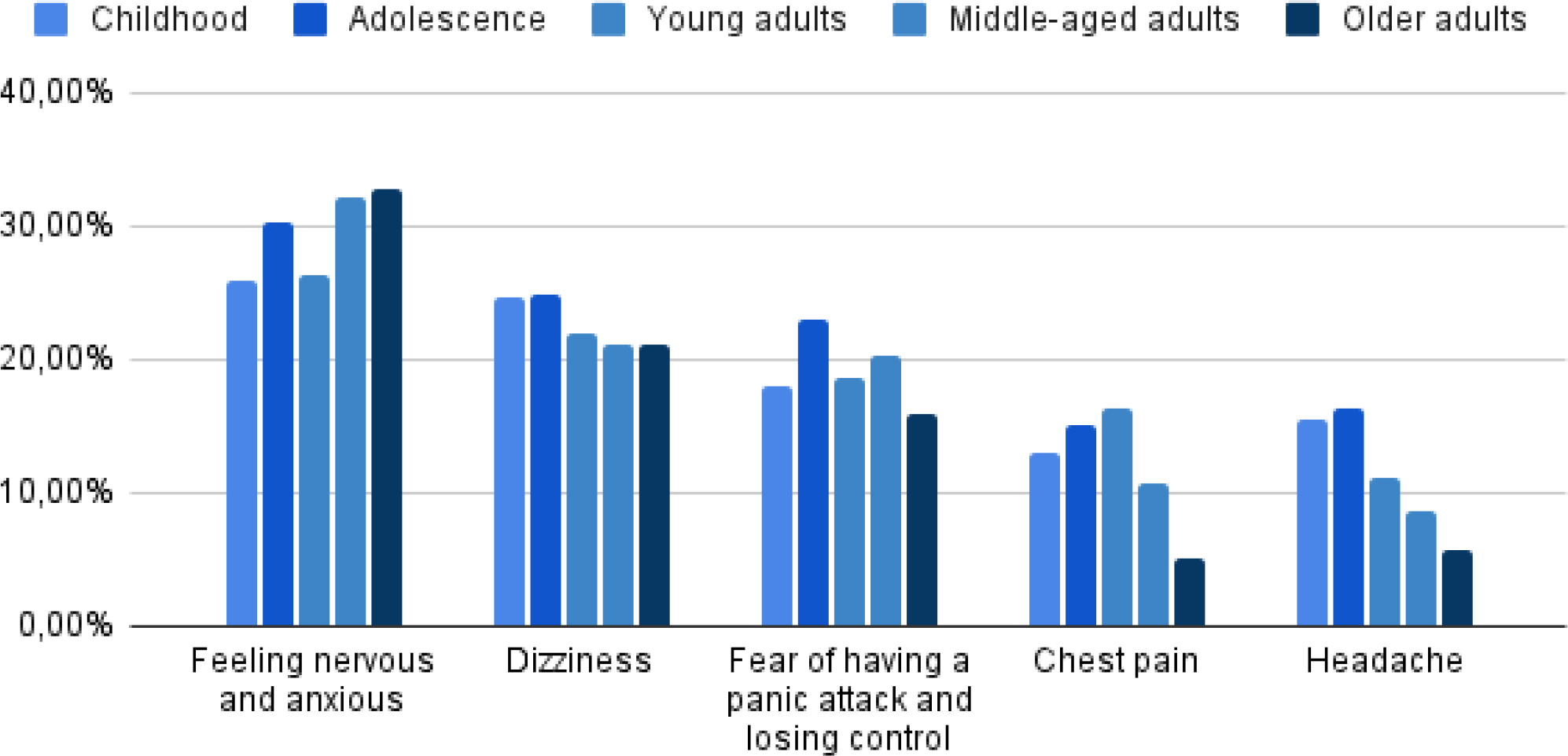
The different reasons for consultation and their distribution according to life stage.

## Discussion

In our study, we observed that it is more common for women to consult for anxiety disorders than for men. This finding is consistent with a study by the Department of Psychology at Boston University, which demonstrated that women have a greater lifetime risk of developing anxiety disorders than men do. Notably, the persistence of AD did not differ between genders^13^.

Women with a diagnosis of AD are more likely to be diagnosed with another disorder, such as bulimia or major depressive disorder, compared to men. Additionally, studies have not shown significant gender differences in the average age of onset of anxiety disorders^13^.

Our findings indicate that young adults were the most frequently diagnosed population with anxiety disorders during the study period. Typically, AD have an onset between the ages of 21 and 35. Some studies have suggested that childhood anxiety symptoms may persist into adulthood, and early identification and intervention may prevent the development of more severe conditions^14^.

A recent study on the prevalence of mental disorders among young Saudis aged 15-30 years old sheds light on the high incidence of anxiety disorders in both young adults and adolescents. This study suggests that increased stress and pressure in modern life, including academic, work, and social demands, may contribute to this trend^15^.

Additionally, hormonal changes and brain development during adolescence and young adulthood can increase susceptibility to AD, while exposure to traumatic events, such as violence or abuse, may increase the risk. However, further research is required to better understand the underlying causes of anxiety disorders in this population^15^. Compounding this issue are barriers to accessing mental health care, such as stigma, a lack of awareness, and inadequate social support.

Based on the data from this study, the most frequent reasons for consultation for these disorders included feeling nervous and anxious, experiencing dizziness, fear of losing control or having panic attacks, chest pain, and headache.

The frequency of feeling nervous and anxious increases in youth and middle-aged adults. Dizziness, on the other hand, was more common among individuals under the age of 18. Fear of panic attacks and headaches were more common in individuals over the age of 18 years, while chest pain was more common in children, adolescents, and young adults.

These physical and psychological symptoms are hallmark features of AD. It is crucial to recognize that anxiety disorders can significantly impair the quality of life of affected individuals and can present in various forms, including panic attacks, specific phobias, generalized anxiety disorders, and others^4^.

As demonstrated in previous research, Mediktor has been recognized as a pre-diagnostic tool that can effectively complement traditional clinical practices. In an analysis of Mediktor’s diagnostic outcomes, the system achieved a diagnostic accuracy rate of 91.3% when the results were analyzed against a list of 10 possible pathologies indicated as diagnoses by Mediktor in an Emergency Department setting. These findings underscore the potential utility of Mediktor as a valuable diagnostic aid that can support clinical decision-making and enhance patient care in various healthcare settings^16^.

In the field of mental health, artificial intelligence (AI) has been a subject of interest in recent years. Technological advances in data collection and analysis allow for greater knowledge and understanding of mental illnesses, such as AD.

AI can help overcome some of the limitations of traditional diagnostic methods, such as lack of objectivity and variability in symptom interpretation, improving diagnostic accuracy, and personalized medical care.

## Conclusion

According to this study, the data observed in Mediktor relates to current scientific evidence. These findings highlight Mediktor’s potential as a technological tool for early detection of anxiety disorders, helping to bridge the gap between patients and healthcare professionals. Therefore, this study emphasizes the importance of artificial intelligence tools to improve the early detection of mental health illnesses.

## Data Availability

All data produced in the present study are available upon reasonable request to the authors

